# The Golden Opportunity or the Cutting Room Floor? Quantifying and Characterizing the Loss and Addition of Social Determinants of Health during Clinician Editing of Ambient AI Documentation

**DOI:** 10.64898/2026.04.20.26351322

**Authors:** Seungjun Kim, Yawen Guo, Sairam Sutari, Emilie Chow, Steven Tam, Danielle Perret, Deepti Pandita, Kai Zheng

**Affiliations:** Department of Informatics, University of California, Irvine, Irvine, CA, USA; Institute for Clinical and Translational Science, University of California, Irvine, Irvine, CA, USA; Department of Medicine, University of California, Irvine, Irvine, CA, USA; Department of Physical Medicine & Rehabilitation, University of California, Irvine, Irvine, CA, USA

## Abstract

Social determinants of health (SDoH) are important for clinical care, but it remains unclear how much AI-captured social context is preserved after clinician editing in ambient documentation workflows. We retrospectively analyzed 75,133 paired ambient AI–drafted and clinician-finalized note sections from ambulatory care at a large academic health system. Using a rule-based NLP pipeline, we extracted 21 SDoH categories and quantified retention, deletion, and addition. SDoH appeared in 25.2% of AI drafts versus 17.2% of final notes. At the mention level, AI captured 29,991 SDoH mentions, of which 45.1% were deleted, 54.9% were retained with clinicians adding 3,583 new mentions. Insurance and marital status were most often deleted, whereas substance use and physical activity were more often retained. Deletion patterns also varied by specialty, supporting the need for specialty-aware ambient AI systems.

## Introduction

Social determinants of health (SDoH) exert significant influence on healthcare utilization, care outcomes, and health equity^1,2^. Comprehensive documentation of SDoH in clinical notes is essential for risk stratification, care coordination, population-health management, and addressing structural drivers of disparity^3^. However, time pressure, competing priorities during the encounter, and perceived irrelevance to the immediate clinical problem may act as challenges to capturing social context^4^. This documentation gap may limit point-of-care decision-making, population health management, and downstream research that depends on reliable social context signals^5,6^.

While the primary objective is to reduce documentation burden and combat clinician burnout, ambient artificial intelligence (AI) documentation softwares offer a golden opportunity to overcome these limitations. By passively listening to patient-clinician conversations and summarizing draft notes in real time, these tools automatically transcribe and synthesize rich social-history content that might otherwise be omitted or abbreviated^7^. This record of the dialogue creates a window into the lived experiences of patients, including employment, housing, family support, substance use, and other SDoH domains^7–9^.

However, the path from an AI-draft to the final clinical record is mediated by the clinician. Previous research has shown that while ambient AI improves work efficiency and reduces documentation burden, clinicians can selectively insert one or more smart note sections into a note and they frequently engage in significant editing of these drafts^10^. Qualitative analyses suggest that these edits are driven by a variety of rationales, ranging from correcting inaccuracies to refining the note for clinical conciseness^10,11^. This human-in-the-loop process introduces a critical tension between the comprehensiveness of what AI captures and the clinician’s perception of clinical relevance. While previous work has characterized the general patterns and rationales of clinician edits^12,13^, the specific impact on SDoH documentation remains unexplored. In particular, it is unclear which categories of social history information remain during the transition from draft to final notes, which might be considered non-essential by the editing clinician, and which SDoH information that were not originally included in AI notes are added.

In this study, we utilize a dataset of approximately 75,000 clinical note sections to quantify the deletion and addition of SDOH information. We compare ambient AI-drafted note sections against the final clinician edited note sections and categorize the added or deleted SDoH items into 21 SDOH categories. By characterizing the patterns of data deletion and addition, we aim to identify which social factors are most vulnerable to deletion and provide insights into how ambient AI systems can be better optimized to preserve essential social context without infringing on clinical utility.

## Methods

### Study design and Data Source

This retrospective, observational study was conducted on ambient AI-drafted and clinician-finalized notes from the University of California, Irvine Health (UCI Health), where a commercial ambient AI documentation tool (Abridge AI, Inc., Pittsburgh, PA, USA)^14^ was piloted and deployed across various ambulatory primary and specialty care clinics. By capturing patient-clinician conversations, the tool automatically drafts clinical note sections that clinicians can selectively insert into a note, including History of Present Illness (HPI), Assessment and Plan (A&P), Physical Exam, and Results. Clinicians then review these drafts before finalizing the electronic health record (EHR) notes. All data processing was conducted within HIPAA-compliant Amazon Web Services (AWS) infrastructure. The study was approved by the University of California, Irvine Institutional Review Board (IRB #7123). Analyses were conducted in Python 3.12 using the pandas and difflib libraries. The final analytic sample consisted of 75,133 note section pairs of AI-draft sections and their corresponding clinician-finalized versions between September 26, 2024, and August 5, 2025.

### SDoH extraction

To prepare the clinical narratives for social determinants of health (SDOH) extraction, we applied a standardized text normalization pipeline to both the AI-drafts and the final clinician-edited notes. Text was lowercased, punctuations such as typographic quotes were normalized, and whitespaces were collapsed.

We developed a rule-based Natural Language Processing (NLP) extraction pipeline involving regular expressions and dictionaries to identify SDoH mentions at the sentence level. The taxonomy mapped to 21 distinct SDOH categories and corresponding sub-categories within each category established in the most recent literature on extracting SDoH from clinical notes^15^, encompassing domains such as housing status, employment, food insecurity, substance use, and demographic identities (Table 1). During this process, a catalog of regular-expression patterns and rules representing each SDoH category and subcategory was constructed. Prior studies that used NLP pipelines based on rules, dictionaries, and regular expression for extracting SDoH from clinical notes were referenced for this construction process^16,17^.

**Table 1.**
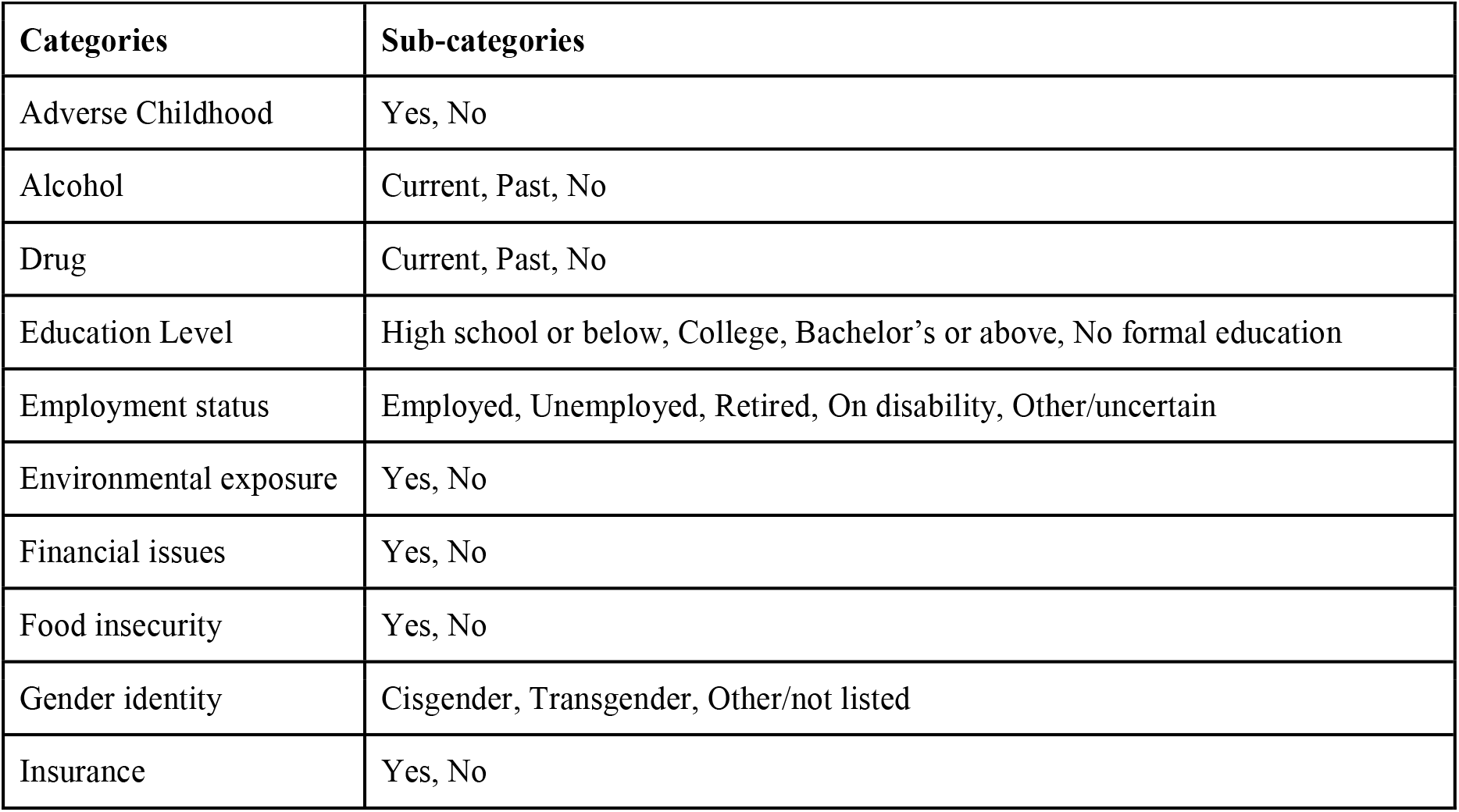
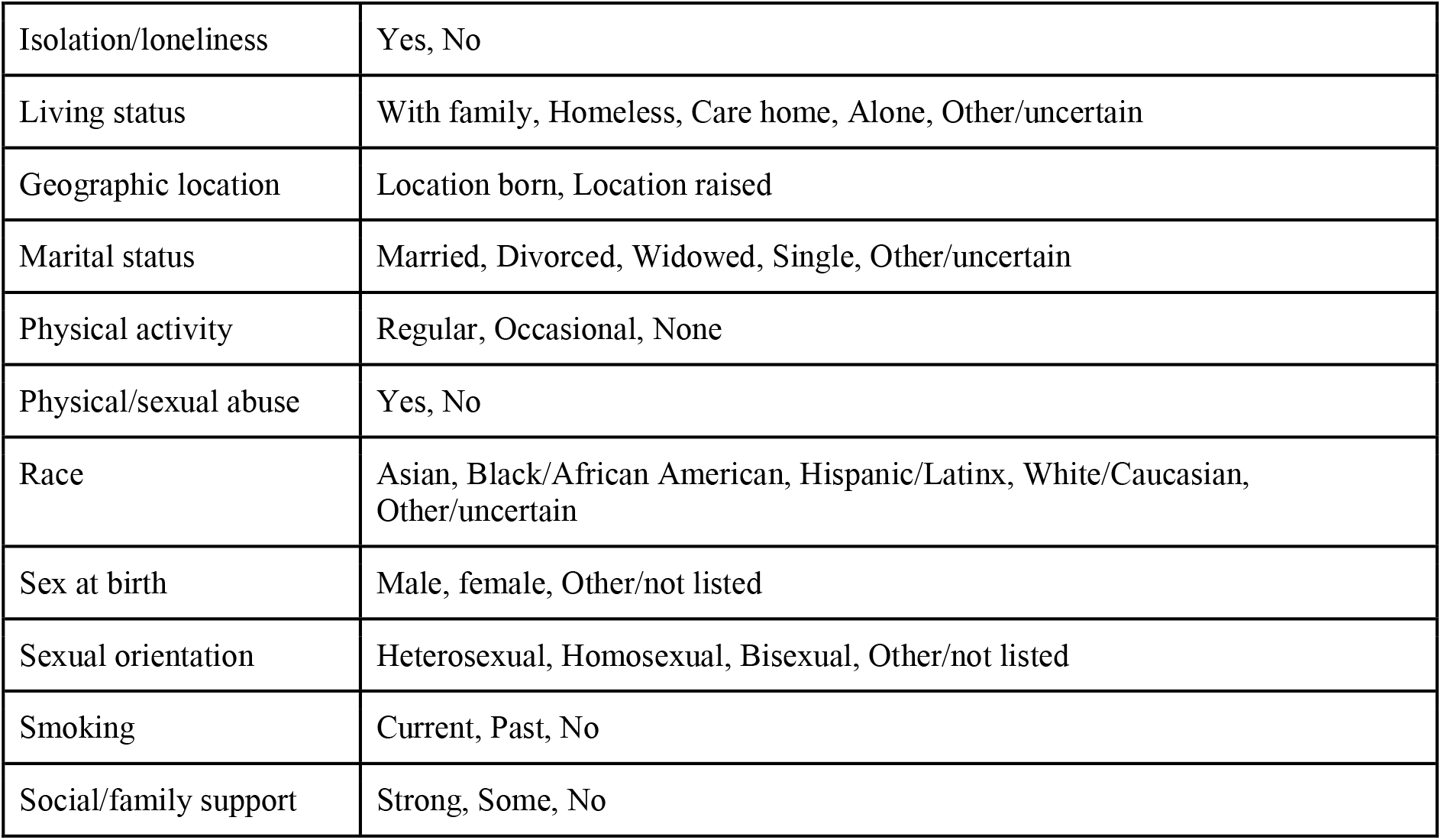
21 SDoH categories with their corresponding sub-categories.

| Categories | Sub-categories |
| --- | --- |
| Adverse Childhood | Yes, No |
| Alcohol | Current, Past, No |
| Drug | Current, Past, No |
| Education Level | High school or below, College, Bachelor's or above, No formal education |
| Employment status | Employed, Unemployed, Retired, On disability, Other/uncertain |
| Environmental exposure | Yes, No |
| Financial issues | Yes, No |
| Food insecurity | Yes, No |
| Gender identity | Cisgender, Transgender, Other/not listed |
| Insurance | Yes, No |
| Isolation/loneliness | Yes, No |
| Living status | With family, Homeless, Care home, Alone, Other/uncertain |
| Geographic location | Location born, Location raised |
| Marital status | Married, Divorced, Widowed, Single, Other/uncertain |
| Physical activity | Regular, Occasional, None |
| Physical/sexual abuse | Yes, No |
| Race | Asian, Black/African American, Hispanic/Latinx, White/Caucasian, Other/uncertain |
| Sex at birth | Male, female, Other/not listed |
| Sexual orientation | Heterosexual, Homosexual, Bisexual, Other/not listed |
| Smoking | Current, Past, No |
| Social/family support | Strong, Some, No |

To enhance precision and reduce false-positive classification in clinical text, the extraction pipeline incorporated 2 rule-based gating mechanisms. First, negative look around filters were used to suppress matches in contexts where candidate terms reflected clinical rather than social-history content. For instance, expressions such as “white blood cell” were excluded from race/ethnicity-related detection, while phrases such as “drug allergy” and “adverse drug reaction” were excluded from illicit drug use-related detection. Second, for demographic domains prone to contextual ambiguity, including race/ethnicity, sexual orientation, and gender identity, the algorithm required co-occurrence of predefined contextual anchors within the same sentence (e.g., “identifies as,” “pronouns,” or “orientation”) to register a valid mention.

### Measuring deletion, and addition of SDoH

To quantify the retention and deletion of SDOH data, we conducted a comparative analysis between the extracted SDOH entities in the AI draft and the finalized note. While token-level Myers diff algorithms were utilized for general string-level edit alignments, semantic SDOH data loss was computed using category-level set differences^18^. Specifically, an SDOH data point was classified as deleted if the (category, subcategory) pair existed in the AI draft but was absent in the finalized text, added if it appeared in the final text but not the draft, and retained if it existed in both versions.

We computed both note-level and mention-level metrics: (1) Proportion of notes containing one or more SDOH element in the AI-draft vs. the final note sections; (2) Proportion of notes with one or more deletions or additions; (3) Total counts and percentages of deleted, and newly added SDOH mentions, stratified by SDoH categories. To understand contextual variations in documentation behavior, these editing patterns of SDOH insertions and deletions were aggregated and stratified across multiple clinical subgroup variables as well, including encounter type, patient class/visit type, visit department, and clinician specialty.

## Results

Among the 75,133 note-section pairs, 18,935 (25.2%) AI drafts and 12,911 (17.18%) final sections contained one or more SDoH items (Table 2). Clinician editing led to deletions of 8,691 note sections (11.57%) with SDoH and additions of 2,617 notes (3.48%) with SDoH.

**Table 2.** Overall SDoH capture and editing (note-level)

| Type of Note Sections and SDoH Edits | Count | % of Note Sections (N=75,133) |
| --- | --- | --- |
| ≥1 SDoH in AI-drafted note section | 18,935 | 25.2% |
| ≥1 SDoH in final note section | 12,911 | 17.18% |
| ≥1 SDoH deletion | 8,691 | 11.57% |
| ≥1 SDoH addition | 2,617 | 3.48% |
| ≥1 SDoH retained | 10,856 | 14.45% |

At the mention-level, ambient AI produced 29,991 SDoH mentions, of which 16,463 (54.9%) were retained and 13,528 (45.1%) were deleted (Table 3). Clinicians added 3,583 new SDoH mentions not present in AI-drafted note sections. The average number of deleted SDoH mentions per affected note was 1.56, suggesting that when deletion occurs, clinicians often remove multiple SDoH items from the same note.

**Table 3.** Overall SDoH capture and editing (mention-level)

| Type of Note Sections and SDoH Edits | Count |
| --- | --- |
| SDoH mentions in AI-drafted note sections | 29,991 |
| SDoH mentions in final note sections | 20,046 |
| Deleted SDoH mentions | 13,528 |
| Added SDoH mentions | 3,583 |
| Retained SDoH mentions | 16,463 |

Deleted SDoH mentions were concentrated in a small set of categories (Table 4). The largest contributors were Insurance (2,960; 21.88% of deletions), Employment status (2,245; 16.60%), Marital status (2,123; 15.69%), and Smoking (1,770; 13.08%), followed by Physical activity, Drug use, and Education level. Added mentions were dominated by the same categories including Marital status (1,150; 32.10% of additions), followed by Insurance (551; 15.38%), Employment status (411; 11.47%), Alcohol use (401; 11.19%), and Smoking (334; 9.32%). This may indicate that while clinicians often remove administrative/demographic content, they also sometimes introduce or correct these domains during finalization.

**Table 4.** Top 10 deleted and added SDoH categories (mention-level)

| Rank | Deletion Category | Deleted mention count (%) | Addition Category | Added mention count (%) |
| --- | --- | --- | --- | --- |
| 1 | Insurance | 2,960 (21.88%) | Marital status | 1,150 (32.10%) |
| 2 | Employment status | 2,245 (16.60%) | Insurance | 551 (15.38%) |
| 3 | Marital status | 2,123 (15.69%) | Employment status | 411 (11.47%) |
| 4 | Smoking | 1,770 (13.08%) | Alcohol use | 401 (11.19%) |
| 5 | Physical activity | 1,143 (8.45%) | Smoking | 334 (9.32%) |
| 6 | Drug use | 864 (6.39%) | Physical activity | 245 (6.84%) |
| 7 | Education level | 863 (6.38%) | Drug use | 149 (4.16%) |
| 8 | Alcohol use | 709 (5.24%) | Living status | 142 (3.96%) |
| 9 | Living status | 578 (4.27%) | Education level | 94 (2.62%) |
| 10 | Financial issues/income | 70 (0.52%) | Physical/sexual abuse | 21 (0.59%) |

To quantify clinician filtering and SDoH retention, we computed retention conditional on SDoH items captured by Ambient AI (e.g., Number of retained SDoH items / Number of SDoH items captured by AI). The clearest pattern emerged among high-volume categories (Table 5). Smoking and Physical activity showed relatively high retention rates (59.43% and 63.31%). Insurance, Employment, and Marital status hovered near 50–53% retention rates, suggesting substantial pruning of administrative/demographic context. Identity-related domains showed higher deletion rates (e.g., Sexual orientation 80% deleted; Race/ethnicity 71.4% deleted) although these categories had low-frequencies and should be interpreted cautiously.

**Table 5.** SDoH retention and deletion rates within SDoH mentions in AI-drafts by major SDoH categories.

| Category | SDoH mentions in AI-drafts | Retained (%) | Deleted (%) | Added |
| --- | --- | --- | --- | --- |
| Insurance | 5,890 | 2,930 (49.75%) | 2,960 (50.25%) | 551 |
| Employment status | 4,600 | 2,355 (51.20%) | 2,245 (48.80%) | 411 |
| Marital status | 4,562 | 2,439 (53.46%) | 2,123 (46.54%) | 1,150 |
| Smoking | 4,363 | 2,593 (59.43%) | 1,770 (40.57%) | 334 |
| Physical activity | 3,115 | 1,972 (63.30%) | 1,143 (36.69%) | 245 |
| Drug use | 2,028 | 1,164 (57.40%) | 864 (42.60%) | 149 |
| Education level | 1,817 | 954 (52.50%) | 863 (47.50%) | 94 |
| Alcohol use | 1,694 | 985 (58.15%) | 709 (41.85%) | 401 |
| Living status | 1,285 | 707 (55.02%) | 578 (44.98%) | 142 |

**Table 6.** Number and proportion of deleted SDoH mentions by top 10 subcategories.

| Rank | Category | Subcategory | Deleted SDoH mention counts<br>(% out of all deleted SdoH mentions) | Category | Subcategory | Added SDoH mention counts<br>(% out of all deleted SdoH mentions) |
| --- | --- | --- | --- | --- | --- | --- |
| 1 | Insurance | Yes | 2,940 (21.73%) | Marital status | Married | 885 (24.70%) |
| 2 | Marital status | Married | 1,827 (13.51%) | Insurance | Yes | 516 (14.40%) |
| 3 | Employment status | Employed | 1,718 (12.70%) | Alcohol use | Current | 358 (9.99%) |
| 4 | Physical activity | Regular | 1,043 (7.71%) | Employment status | Employed | 259 (7.23%) |
| 5 | Smoking | Current | 989 (7.31%) | Marital status | Single | 258 (7.20%) |
| 6 | Drug use | Current | 810 (5.99%) | Smoking | Current | 223 (6.22%) |
| 7 | Education level | College | 634 (4.69%) | Physical activity | Regular | 214 (5.97%) |
| 8 | Smoking | No | 587 (4.34%) | Drug use | Current | 140 (3.91%) |
| 9 | Alcohol use | Current | 586 (4.33%) | Employment status | Retired | 91 (2.54%) |
| 10 | Living status | With family | 344 (2.54%) | Living status | With family | 64 (1.79%) |

Subcategory analyses revealed that the single largest deleted subcategory was [Insurance: Yes] (2,940) (Table 4). Other high-frequency deletions included [Marital status: Married] (1,827; 13.51%) and [Employment status: Employed] (1,718; 12.7%). Among health behaviors, clinicians most commonly deleted [Smoking: Current] (989; 7.31%) and [Drug use: Current] (810; 5.99%), indicating that even clinically relevant behaviors are pruned in many notes, likely reflecting encounter-specific relevance judgments or redundancy elsewhere in the chart.

SDoH deletion and retention varied widely by clinician specialty, supporting the interpretation that documentation norms are context dependent. Otolaryngology showed high SDoH deletion prevalence (45.99% of notes with deletion) and very high deletion intensity (94.29 deletions per 100 SDoH mentions in AI-drafts). Internal Medicine showed substantial deletion prevalence (21.43%) and high deletion intensity (62.81 per 100 SDoH mentions in AI drafts), consistent with frequent editing of administratively redundant SDoH. Emergency Medicine displayed high deletion intensity (90.74 per 100 SDoH mentions in AI drafts) with moderate deletion prevalence (14.81%), consistent with acute-problem focus and time-pressured documentation.

Department-level outputs similarly displayed heterogeneous patterns, indicating that clinical setting influences SDoH curation. For example, several internal medicine and primary care departments exhibited elevated deletion intensity (e.g., UCI PAV3 INTERNAL MED: 74.20 deletions per 100 SDoH mentions in AI drafts), suggesting that when ambient AI captures social context in these settings, clinicians frequently prune it to align with note conventions and concision.

## Discussion

This study quantifies clinicians’ curation of AI-captured social context in ambient AI documentation workflows. Across 75,133 ambient AI note-section pairs, ambient AI surfaced SDoH in 25.2% of drafts, but only 17.2% of final notes contained SDoH, highlighting a meaningful gap between conversationally captured social context and what clinicians ultimately record. More specifically, 45.1% of SDoH mentions in AI-drafted note sections were deleted during editing, underscoring a substantial filtering process. Concurrently, clinicians introduced 3,583 new SDoH mentions, showing an active edit process in shaping the final record.

Moreover, deletions were highly concentrated in certain SDoH categories. Insurance, employment status, and marital status collectively comprised more than half of all deletions. Retention analysis further demonstrated that behaviors such as smoking and alcohol or drug use were relatively more likely to persist, while administrative and demographic domains were pruned heavily. Subcategory analyses revealed that deletions frequently targeted normative and administrative attributes (e.g., insured, employed, married, regular exercise) and common living arrangements (e.g., living with family), suggesting the editing process is not merely correcting errors but actively shaping the informational content of the medical record^10,19^.

These patterns may reflect a two-factor model of clinician curation: clinical relevance and redundancy with information documented elsewhere. Clinicians were more likely to retain SDoH that directly informed risk assessment or care planning (e.g., smoking, substance use, and some living status indicators). By contrast, they more often removed information that may already exist in structured EHR fields, administrative metadata, or other standardized sections, and thus may have been considered unnecessary to repeat in the narrative note^20,21^. This category may include insurance, marital status, employment, and some identity-related variables (e.g., sexual orientation, race/ethnicity, birthplace). Accordingly, higher deletion rates for these domains should be interpreted cautiously and not as evidence that such information is unimportant.

These findings point to a documentation gap between what ambient AI captures from clinical conversations and what clinicians ultimately choose to record in the EHR. The pattern appears to reflect clinician judgments about relevance, note utility, redundancy, and documentation norms. This highlights an important opportunity for ambient AI design. Systems should better align with clinician documentation needs by improving how socially and contextually relevant information is categorized and prioritized. Some detailed social factors may be important to retain in structured social history forms without being kept for the note itself. Future ambient AI systems should therefore support both concise, clinically useful notes and comprehensive capture of SDoH by helping route information to the most appropriate location in the record.

Subgroup analyses demonstrated contextual variability, with deletion rates and intensity differing by clinician specialty and department. For instance, otolaryngology and emergency medicine exhibited aggressive pruning (e.g., 94.3 and 90.7 deletions per 100 SDoH mentions in AI-drafted note sections, respectively), likely reflecting acute-focus priorities, whereas internal medicine showed more moderate but still substantial filtering (62.8 per 100). Such heterogeneity implies that SDoH documentation is not uniform but influenced by specialty-specific workflows, encounter acuity, and documentation conventions. This finding calls for a specialty-specific AI design approach and advocates for adaptive systems tailored to clinical contexts.

Several limitations warrant consideration. First, this study was conducted at a single institution using one ambient AI platform, which may limit generalizability across health systems, specialties, documentation cultures, and products. Multi-site and multi-vendor studies are needed to determine whether the observed SDoH retention, addition, and deletion patterns hold across settings. Second, our rule-based extraction pipeline was intentionally optimized for precision, which likely reduced false positives but may also have underestimated the full volume of SDoH content. Future work should evaluate hybrid approaches that combine rule-based methods with machine learning- or large language model-assisted extraction to improve recall while preserving acceptable precision^15^. Third, we did not assess the clinical appropriateness of individual deletions or whether some information was relocated to other note sections, so removed content should not automatically be interpreted as meaningful loss. Future studies should incorporate richer encounter context and section-level analyses to better distinguish justified editing, relocation, and true omission. Finally, some low-frequency SDoH domains were too sparse to support strong inferences, highlighting the need for larger or more targeted datasets.

## Conclusion

This study reveals that while ambient AI holds promise for enriching clinical documentation with comprehensive SDoH insights from patient-clinician dialogues, clinician editing introduces significant filtering, with 45.1% of SDoH mentions in AI-drafted note sections deleted and a bias toward retaining risk-related details over neutral or administrative ones. These patterns, varying by specialty and department, underscore the need for context-aware AI designs that balance comprehensiveness with clinical utility. By optimizing ambient AI to align with clinician workflows and ethical considerations, we can better preserve essential social context, mitigate documentation biases, and ultimately advance health equity and holistic patient care. Future efforts should focus on interdisciplinary collaborations to refine these technologies, ensuring they serve as tools for empowerment rather than sources of unintended data loss.

## Data Availability

All data produced in the present study are available upon reasonable request to the authors

